# Delayed-Phase Thrombocytopenia in Patients of Coronavirus Disease 2019 (COVID-19)

**DOI:** 10.1101/2020.04.11.20059170

**Authors:** Wanxin Chen, Ziping Li, Bohan Yang, Ping Wang, Qiong Zhou, Jianhua Zhu, Xuexing Chen, Peng Yang, Hao Zhou

## Abstract

The pandemic COVID-19 pneumonia has engulfed the entire world. Hematopoietic system can also be affected by COVID-19. Thrombocytopenia at admission was prevalent, while late-phase or delayed-phase thrombocytopenia is obscure. This retrospective single-center case series analyzed patients with COVID-19 at the Union Hospital, Wuhan, China, from January 25th to March 9th, 2020. Analysis began on March 11th, 2020. COVID-19 associated delayed-phase thrombocytopenia was occurred in 11.8% percent of enrolled patients. The delayed-phase thrombocytopenia in COVID-19 is prone to develop in elderly patients or patients with low lymphocyte count on admission. The delayed-phase thrombocytopenia is significantly associated with increased length of hospital stay and higher ICU admission rate. Delayed-phase nadir platelet counts demonstrated a high and significantly negative linear correlation with B cell percentages and serum IL-6 levels. We also presented bone marrow aspiration pathology of three patients with delayed-phase thrombocytopenia, showing impaired maturation of megakaryocytes. We speculated that the delayed-phase platelet destruction might be mediated by antibodies, and suggest immunoregulatory treatment in severe patients to improve outcomes. Besides, clinicians need to pay attention to the delayed-phase thrombocytopenia especially at 3-4 weeks after symptom onset.

## INTRODUCTION

In December 2019, the outbreak of severe viral pneumonia of unknown origin was initially reported in Wuhan, China. A novel coronavirus was identified as the causative agent and was firstly named as 2019 novel coronavirus (2019-nCoV). This disease was eventually termed coronavirus disease 2019 (COVID-19) and the virus severe acute respiratory syndrome coronavirus 2 (SARS-CoV-2) [1, 2]. The pandemic COVID-19 has engulfed the entire world with an estimated mortality rate of 4.8% [3].

Common symptoms of COVID-19 include fever, cough, and shortness of breath, since lung is the major target of SARS-CoV-2. Muscle pain, sputum production, diarrhea, sore throat and loss of taste and/or loss of smell are possible but less common. Cardiac and neurologic complications were also found in COVID-19. These extra-pulmonary manifestations imply diverse target organs in addition to lung. Hematopoietic system can also be affected by COVID-19. A multicenter study by our hospital and other centers has demonstrated that on admission, lymphocytopenia was present in 83.2% of the patients, thrombocytopenia in 36.2%, and leukopenia in 33.7% [4].

With increased hospitalization capacity and prolonged isolation period, we are now able to study the disease longitudinally apart from cross-sectionally on admission. In particular, our initial observations showed there were sudden dramatic decline in platelet count in several COVID-19 patients without evidence of other coagulation abnormalities, which happened three weeks or more after symptoms onset. COVID-19 related early-phase thrombocytopenia was prevalent [5], while late-phase or delayed-phase thrombocytopenia is obscure.

In the current study, we reported the incidence, characteristics, and outcomes of patients with delayed-phase thrombocytopenia. We also presented bone marrow aspiration pathology of three patients with delayed-phase thrombocytopenia.

## METHODS

### Study design and participants

This was a retrospective, descriptive, longitudinal study, conducted from January 25th to March 9th, 2020. Analysis began on March 11th, 2020. All patients were recruited from the Union Hospital, Tongji Medical College, Huazhong University of Science and Technology, Wuhan, China. The study was performed in accordance to the principles of the Declaration of Helsinki and was approved by the Research Ethics Committee of Union Hospital, Tongji Medical College, Huazhong University of Science and Technology (No. 2020-0079-1).

There were two inclusion criteria for this study: (1) each patient was confirmed by real-time reverse transcription PCR (real-time RT-PCR) and were diagnosed as having COVID-19 according to WHO interim guidance [6]; (2) all patients underwent chest computerized tomography (CT) and complete panel of routine laboratory tests, including compete blood count, blood biochemistry, blood coagulation function, test of key inflammatory cytokines (IL-4, IL-6, IL-10, IFN-γ, TNF-α), and lymphocytes subset analysis. The exclusion criteria were: (1) monitoring period less than 21 days from symptom onset; (2) autoimmune or hematological disease history; (3) previous human immunodeficiency virus (HIV), hepatitis B or hepatitis C infection; (4) dialysis patients. All patients will be screened for the inclusion and exclusion criteria described above.

### Procedures

The demographics data, clinical characteristics, laboratory data, chest CT scan findings, treatment programs, and outcomes were obtained from patients’ medical records. Clinical outcomes were followed up to March 30th, 2020. Any missing or uncertain records were collected and clarified through direct communication with involved health-care providers and their families. The data were reviewed by a trained team of physicians. All data were separately extracted by two authors (Peng Yang and Hao Zhou).

We collected data on age, gender, respiratory rate, smoking and comorbidities (Hepatitis B, Hepatitis C, HIV, hepatosplenomegaly, hematological system disease, rheumatic immune system disease, cardio cerebrovascular disease, endocrine system disease, respiratory system disease, digestive system disease, nervous system disease and malignant tumor), symptoms from onset to hospital admission (fever, cough, dyspnea, pharyngalgia, diarrhea, anorexia, abdominal pain, palpitation, hypodynamia, paresthesia, myalgia and dizziness), laboratory values on admission (blood routine, blood coagulation function, blood biochemistry, C-reactive protein, inflammatory cytokines and lymphocyte Subsets), treatment (antiviral agents, antibacterial agents, antifungal agents, corticosteroids, inhaled interferon-α, immunoglobulin and oxygen therapy), as well as living status. Furthermore, we collected the continuous monitored laboratory data including blood routine, blood coagulation function, inflammatory cytokines (IL-4, IL-6, IL-10, IFN-γ, TNF-α), and lymphocytes subset analysis. For some patients, the SARS-CoV-2 IgG antibody results were recorded when available.

The date of disease onset was defined as the day when the symptom was noticed. The severity of COVID-19 was defined according to the diagnostic criteria of “COVID-19 diagnosis and treatment Plan (trial seventh edition)” issued by the National Health Commission of the People’s Republic of China [7]. Throat swab samples were collected and placed into a collection tube containing preservation solution for the virus [8]. SARS-CoV-2 was confirmed by real-time RT-PCR assay with SARS-CoV-2 nucleic acid detection kit (Shanghai bio-germ Medical Technology, Shanghai, China). The IgG antibody were detected with SARS-CoV-2 IgG antibody detection kit (YHLO biotechnology, Shenzhen, China).

### Study outcomes

We extracted the demographic data, clinical characteristics including respiratory and extra-respiratory symptoms on admission, comorbidities, laboratory data, treatment programs, and clinical outcomes (remained in hospital isolation ward, in ICU, discharged, or died). This outcome category was used in a previous COVID-19 study [9].

### Statistical analysis

Descriptive data were presented as means (± standard deviation [SD]) for normally distributed continuous variables and as medians with interquartile range (IQR) for non-normally distributed data. Categorical variables were presented as percentages. Two independent samples were tested by T-test; the analysis of variance or Kruskal-Wallis rank sum test was applied for comparison between multiple groups. Proportions for categorical variables were compared using the χ2 test, although the Fisher exact test was used when data were limited. The Pearson correlation coefficient and Spearman rank correlation coefficient were used for liner correlation analysis. All statistical analysis was performed using SPSS v.19.0 (IBM Corp., Armonk, NY, USA). A two-tailed *P value* <0.05 was considered statistically significant.

## RESULTS

### Patient flow and baseline characteristics

This resulted in an analyzable population of 271 COVID-19 positive patients. The last follow-up at the time of writing this study was March 30th, 2020. Patient baseline characteristics are shown in Table 1. The average age was 57.4 years (SD = 14.3), including 145 men and 126 women. The average time from symptom onset to hospital admission was 6.65 days (SD = 2.78). There were 75 (27.7%) patients with cardiovascular diseases, 35 (12.9%) patients with endocrine system diseases, and others with respiratory diseases (11 [4.1%]), malignant tumors (9 [3.3%]) and other disorders as shown in Table 1. After admission, there were 246 (90.8%) patients receiving antiviral treatment, 176 (64.9%) on antibiotics, 115 (42.4%) on glucocorticoids, 117 (43.2%) on inhaled interferon-α, and 65 (24.0%) on intravenous immunoglobulin. Additionally, 28 patients were transferred to the ICU (10.3%), 60 were still remaining in hospital isolation ward at the time this study was completed (22.1%), 177 were discharged (65.3%), and 6 (2.2%) died. Among the discharged patients, the average hospital isolation ward stay was 31.35 days (SD = 11.20).

### Incidence and outcomes of patients with delayed-phase thrombocytopenia

Based on the presumed viremia phase after SARS-CoV-2 infection [10] and our preliminary clinical observations, we propose to set 14 days as the rough while reasonable and feasible cutoff value. Therefore, delayed-phase thrombocytopenia is defined as thrombocytopenia begins or worsens after 14 days post symptoms appearance. We found 32 patients (11.8 %) developed delayed-phase thrombocytopenia (Table1). The median time for delayed-phase thrombocytopenia nadir appeared at 28.3 days, from illness onset. The median duration time for delayed-phase thrombocytopenia was 4.32 (SD = 2.15). We also found that the mean platelet count at nadir to be 86.0 × 10^9^/L (SD = 37.48). Delayed-phase thrombocytopenia is more prevalent in elderly person (71.9% in over 60-years-old patients). The clinical outcomes (in hospital isolation ward, in ICU, discharged, or died) for these 32 patients with delayed-phase thrombocytopenia were 7 patients remained in hospital isolation ward, 16 patients discharged, 7 patients in ICU, and 2 patients died. The representative platelet count curves of COVID-19 patients were shown in Figure 1.

**Figure 1.**
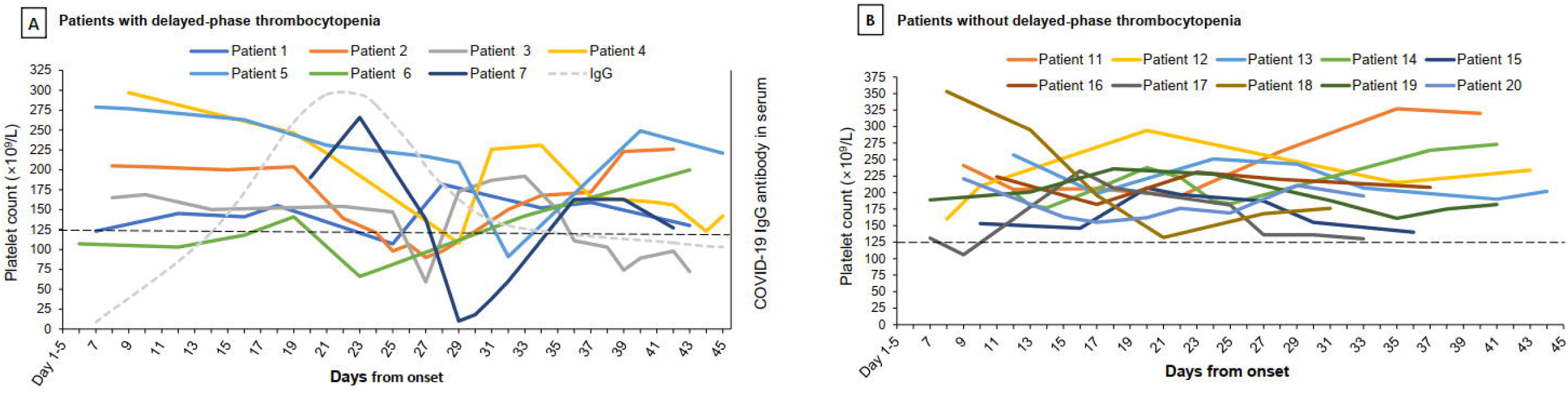
Dynamic changes of platelet count in COVID-19 patients. The day of symptoms onset = Day 1. **A**. The variation tendency of seven representative patients with delayed-phase thrombocytopenia. Each patient is presented in a distinct color. Gray dashed line: COVID-19 IgG production curve. **B**. The variation tendency of ten representative patients without delayed-phase thrombocytopenia.

### Laboratory findings of patients with COVID-19 on admission

The delayed-phase thrombocytopenia cases demonstrated lower lymphocyte count (0.89±0.52, *P* = 0.000) and higher levels of lactate dehydrogenase (334.25±154.51 U/L, *P* = 0.013), comparing with the group without delayed-phase thrombocytopenia (Table 2). Meanwhile, the level of the lymphocyte subsets including CD3^+^ T (69.43±10.33%), CD4^+^ T (38.79±12.45%) and B cells (17.28±12.22%) showed significant alterations from their counterpart. A bunch of key inflammatory cytokines was also changed in delayed-phase thrombocytopenia patients, including IL-4, IL-6 and TNF-α. Notwithstanding, the platelet to lymphocyte ration (PLR) showed no discrepancy between patients with and without delayed-phase thrombocytopenia. Furthermore, there were no significant differences in the levels of white blood cell count, platelet count and coagulation indicators between patients with and without delayed-phase thrombocytopenia on admission.

### Laboratory results time-correlated with delayed-phase thrombocytopenia

We conducted time-correlated data analysis for laboratory changes, underlining cytokine and lymphocyte immunity (Table 3). We reviewed the inflammatory cytokines results at around the time delayed-phase thrombocytopenia occurred. The results showed around the platelet count nadir time point, the median level of IL-4 was 15.90±3.77 pg/ml, IL-6 63.22±76.48 pg/ml, and IL-10 12.11±9.57 pg/ml, all significantly higher than the upper limit of the normal value. The lymphocyte subset, monitored at delayed-phase plate count nadir time, showed that the percentage of CD4^+^ T cells was 51.93±14.33 and B cells was 19.76±12.08, both above the average normal value. Delayed-phase nadir platelet counts demonstrated a high and significantly negative linear correlation with B cell percentages (β = –0.562, *P* < 0.001) and serum IL-6 levels (β = –0.413, *P* < 0.001) (Figure 2).

**Figure 2.**
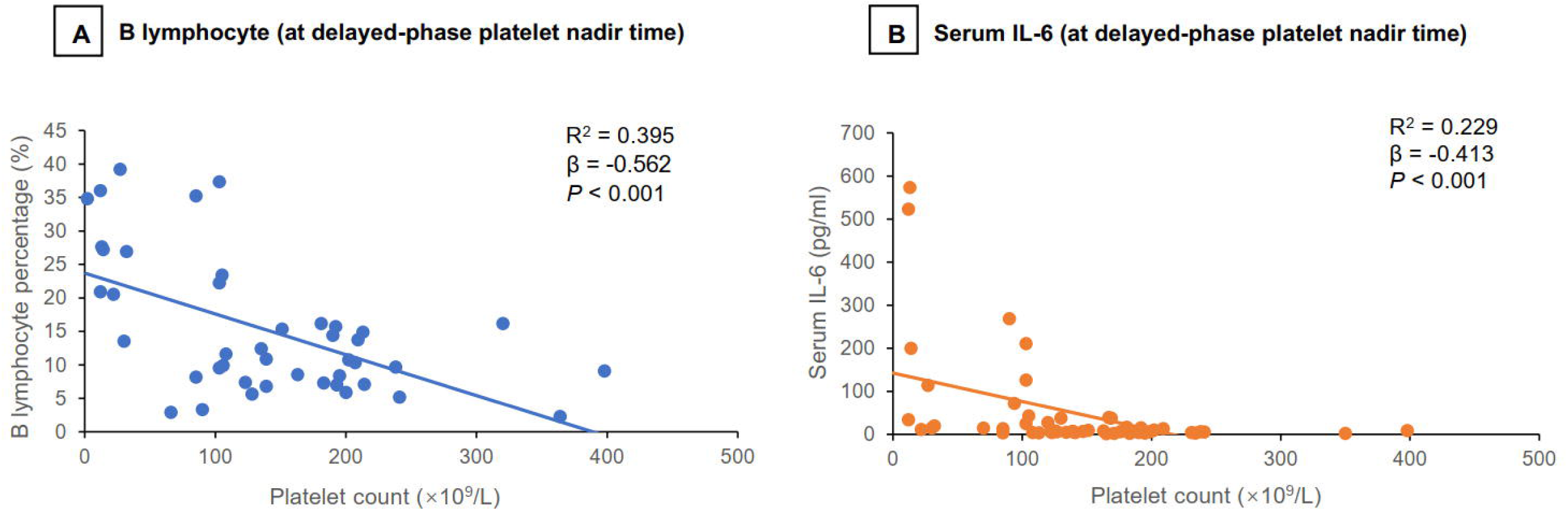
Correlation between B lymphocyte percentage and serum interleukin 6 (IL-6) with platelet count (at the delayed-phase platelet nadir time point).

### Bone marrow aspiration analysis of three cases

We specifically studied three patients underwent bone marrow aspiration in the Union Hospital. All these three patients developed rapid and dramatic decline in platelet count at delayed phase, without evidence of other coagulation abnormalities. The pathology results from each of the three patients shared common features: (1) bone marrow showed no obvious abnormities in the myeloid or the erythroid cells; (2) number of atypical or reactive lymphocytes increased; (3) maturation of megakaryocyte was impaired, the mature platelet-producing megakaryocyte were rare, and most megakaryocytes were immature granular megakaryocytes. Two patients’ bone marrow smear images were presented in Figure 3. In Figure 3E, the mature platelet-producing megakaryocyte was attached by three platelets (one large and the other two small). This might suggest body compensate by making abnormally large platelets.

**Figure 3.**
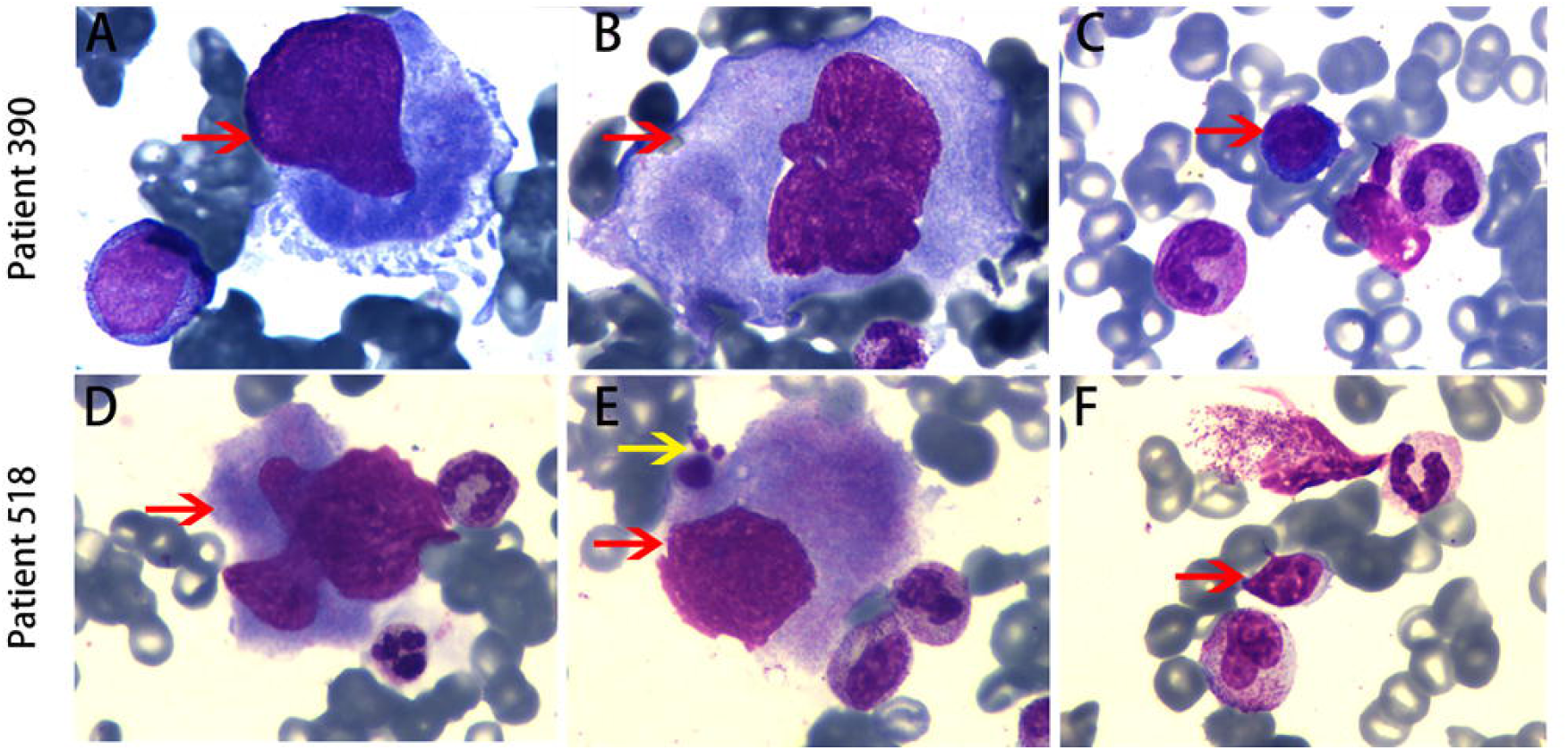
Wright’s stained bone marrow aspirate smears from two patients. **A**. red arrow: granular megakaryocyte (×1000); **B**. red arrow: granular megakaryocyte (×1000); **C**. red arrow: atypical (reactive) lymphocyte (x 400); **D**. red arrow: granular megakaryocyte (×1000); **E**. red arrow: platelet-producing megakaryocyte, yellow arrow: one large and two small platelets (×1000); **F**. red arrow: mature lymphocyte (x400).

## DISCUSSION

The present study was conducted by reviewing the medical records of patients with COVID-19 from January 25th to March 9th, 2020, in a heavily affected hospital during the initial outbreak in China. We found that COVID-19 associated delayed-phase thrombocytopenia was occurred in 11.8% percent of enrolled patients. The delayed-phase thrombocytopenia in COVID-19 is prone to develop in elderly patients or patients with low lymphocyte count on admission. The delayed-phase thrombocytopenia is significantly associated with increased length of hospital stay and higher ICU admission rate.

We began our study based on clinical observation that occasionally several patients developed rapid platelet count decrease after more than 2 weeks of treatment, without evidence of other coagulation abnormalities. Rare cases even suffered severe delayed-phase thrombocytopenia (platelet < 10 × 10^9^/L). Considering natural course of viral infection and treatment outcome of patients, 3 weeks after symptoms onset, most of COVID-19 patients were in the recovery stage [11]. The paradox of delayed-phase thrombocytopenia prompted us to investigate its prevalence and characteristics. With expanded hospitalization capacity and prolonged isolation period, patients can be admitted timely and laboratory results were well followed-up. These enabled us to conduct a longitudinal study on dynamic changes of platelet count.

Our study showed delayed-phase thrombocytopenia is prevalent in the enrolled patients. Thrombocytopenia at admission is frequent in COVID-19 and other virus infection diseases [12-14]. Previous reported contagious coronavirus cases of severe acute respiratory syndrome (SARS) and Middle East respiratory syndrome (MERS) suggested that thrombocytopenia is common in both SARS and MERS patients [15, 16]. However, the viral infection associated delayed-phase thrombocytopenia has merely been reported as rare case reports without enough patients for statistical analysis [17, 18].

To clarify the pathogenesis of the delayed-phase thrombocytopenia, we screened laboratory results, underlining key cytokines and lymphocyte immunity. Our results suggest that IL-6, a strong inflammatory cytokine, was negatively correlated with the delayed-phase platelet count decline. This indicated that IL-6 might be an active player in the delayed-phase platelet decline. Notably, our time-correlated lymphocyte subset analysis showed B cells percentage was negatively associated with the delayed-phase thrombocytopenia. Since antibody production by B cells is crucial in virus protection, this result highlight an important role that antibodies play in the delayed-phase platelet decrease. We also found most of the delayed-phase thrombocytopenia lasted less than 7 days, implying the delayed-phase thrombocytopenia is transient.

Furthermore, we sought to obtain evidence from bone marrow. Bone marrow aspiration analysis help to recognize possible pathogenesis in the hematopoietic tissue. All bone marrow aspiration pathology showed common features as impeded megakaryocyte maturation, and mature platelet-producing megakaryocyte were rare (less than 5 in the bone marrow smear). These bone marrow features were as similar as that in immune thrombocytopenia. Impaired megakaryocyte maturation and insufficient platelet production participate in the pathogenesis of immune thrombocytopenia [19]. At the time-point of platelet count nadir, these three patients had been COVID-19 nucleic acid negative twice (suggesting virus been deleted or suppressed), and their IgG antibody were positive (suggesting protective antibody producing). Based on the antibody production curve after SARS-CoV-2 infection [20], we speculated that at the delayed-phase platelet destruction is mediated by antibodies.

Virus associated immune thrombocytopenia may include several mechanisms [21]. Firstly, a virus-induced change in the host’s immune system by polyclonal B cell activation or release of cytokines. Secondly, production of autoantibodies against platelet glycoproteins induced by the modification of platelet surface proteins by virus infection. Thirdly, cross-reaction of the virus protein-directed antibodies with platelet glycoproteins. Fourthly, virus infected megakaryocyte sheds platelets that presenting viral antigens, so anti-viral antibodies attack against platelets. Animal models with Rauscher virus infection showed viral gp70 antigen expression on platelets correlated with the development of delayed-phase thrombocytopenia [22]. On this ground, treatment using immunoregulatory agents (including intravenous immunoglobulin [IVIG] and dexamethasone) is reasonable and necessary. IVIG was administered to all these three patients thereafter and proved to be effective in 2 to 3 days. A Zika virus-infected patient suffered severe thrombocytopenia 29 days after illness onset. She was treated as presumed immune-mediated thrombocytopenia with IVIG and her platelets recovered in a few days [18].

This study has several limitations. First, because our analysis was based on a retrospective study with a relatively small sample, which could cause biases and limit the reliability of our results. Second, the laboratory data were not monitored by strictly fixed time interval due to rapidly evolving epidemic. This might lead to omission of data at the changing time point. Third, our hospital-based study no doubt missed patients who were mild cases and who were treated at home, and our study cohort may represent the more severe end of COVID-19. Fourth, multivariate regression analysis is recommended to recognize the risk factors and the individual odds ratio. Further study integrating more clinical characters is needed to draw a more complete conclusion. Fifth, because several patients were still hospitalized and information regarding clinical outcomes was unavailable at the time of this writing, the relationship between these patients’ prognosis and delayed-phase thrombocytopenia remains to be investigated.

## CONCLUSIONS

In summary, we have demonstrated prevalence of delayed-phase thrombocytopenia in COVID-19 patients. In rare cases, patients even suffered life-threating thrombocytopenia. Compared to those without delayed-phase thrombocytopenia, COVID-19 patients with delayed-phase thrombocytopenia have a longer hospital stay time and a worse clinical outcome. The present descriptive data suggested an association between delayed-phase thrombocytopenia and antiviral immune, which highlight rationality for immunoregulatory treatment. Our results encourage further research evaluating the prevalence, incidence, predictors, and outcomes of delayed-phase thrombocytopenia in this ongoing pandemic. Meanwhile, doctors need to pay attention to the delayed-phase thrombocytopenia especially at 3-4 weeks after symptom onset, which is the median time for delayed-phase thrombocytopenia occurrence. This knowledge may help doctors to timely apply immunoregulatory treatment, and thus improve outcomes.

## Data Availability

Some or all data during the study are available from the corresponding author by request, under the permission of Ethic Committee and patients.

## Acknowledgements

We acknowledge all health care workers involved in the diagnosis and treatment of patients at Union Hospital.

## Ethical Approval

This study was approved by the Research Ethics Committee of Union Hospital, Tongji Medical College, Huazhong University of Science and Technology.

